# A Bibliometric Analysis of Global Pediatric Emergency Medicine Research Networks

**DOI:** 10.1101/2021.04.14.21255485

**Authors:** Michael J Barrett, Stuart Dalziel, Mark Lyttle, Ronan O’Sullivan, for the Pediatric Emergency Research Networks (PERN)

## Abstract

**Objective:** During the last three decades newly formed pediatric emergency medicine (PEM) research networks have been publishing research. A desire of these networks is to produce and disseminate research to improve patient health and outcomes. To quantitatively analyze and compare the literature by PEM research networks globally through numeric and visual bibliometrics.

**Methods:** A bibliometric analysis of articles published from 1994 to 2019 (26 years) by authors from PEM research networks globally were retrieved using PubMed^®^, Web of Science™ (Thompson Reuters) and accessing individual research network databases. Bibliometric analysis was performed utilizing Web of Science™, VOSviewer and Dimensions. Research was quantified to ascertain the number of articles, related articles, citations and Altmetric attention score.

**Results:** A total of 493 articles were published across nine research networks in three decades. Pediatric Emergency Care Applied Research Network (PECARN) produced the most articles, citations, and h-index of all networks. We identified three main groupings of productive authors across the networks who collaborate globally. The gender of the first author was female in 46% of publications and the corresponding author(s) was female in 45%. A non-significant moderate positive correlation between the number of years publishing and the number of publications was identified. There was non-significant moderate negative association between the number of countries in a network and total publications per annum.

**Conclusions:** This study is the first bibliometric analysis of publications from PEM research networks that collaborate globally. The gender gap in first authorship compared to high impact medical journals and high impact emergency medicine journals is narrower. Exploring the relationships of numerical bibliometric indicators and visualizations of productivity will benefit the understanding of the generation, reach and dissemination of PEM research within the global research community.

## Introduction

Twenty seven years has passed since the first publication by a pediatric emergency medicine (PEM) research network (PEM Collaborative Research Committee (PEM CRC) of the American Academy of Pediatrics). ^1^ In the intervening years eight PEM research networks have united as a global body, the Pediatric Emergency Research Network (PERN), which consists of representatives from four of the six World Health Organization (WHO) regions. The desire of these networks is to produce and disseminate high-quality research to improve health and outcomes of acutely ill and injured children and youths throughout the world. ^2-8^

The global proliferation of research to massive proportions has expanded evidence, but has simultaneously made it difficult to gain oversight, by traditional methods, of the architecture of relevant research to cohere what is published on a given topic or specialty of interest. ^9,10^ Bibliometrics quantifies data for scientific output and has become increasingly important and valuable to researchers, research networks, investing stakeholders, funding agencies and policy makers. ^9,11^ The production of quantitative metrics can challenge bias tendencies in peer review and can facilitate further deliberation on the methods of achieving and optimizing the research missions of the PEM research networks.^11^

The aim of this study is to describe and analyze the literature by global PEM research networks through numeric and visual bibliometrics.

## Methods

### PEM Research Networks

The networks included are Network for Research and Development of PEM in Latin America (Red de Investigacion y Desarrollo de la Emergencia Pediatrica Latinoamericana, RIDEPLA), Research in European Pediatric Emergency Medicine (REPEM), Spanish Pediatric Emergency Research Group (RISeuP/SPERG), Pediatric Emergency Care Applied Research Network (PECARN), Pediatric Emergency Medicine Collaborative Research Committee of the American Academy of Pediatrics (PEM CRC), Pediatric Emergency Research Canada (PERC), Paediatric Research in Emergency Departments International Collaborative (PREDICT) and Paediatric Emergency Research in the United Kingdom and Ireland (PERUKI). Each of these networks are affiliated with PERN.

### Search Strategy/Data Mining

Publications were retrieved from Web of Science™ (Thompson Reuters, Toronto, Canada), Medline^®^/PubMed^®^ and cross referenced from each Research Network (Table 1). A preliminary search of network publications identified 1994 as the first year of publication, which established the timeframe for the analysis of 1994-2019 (Last search 9 March 2020). Publications were identified by searching for the individual research network in the Authorship, Corporate Authorship, Abstract, or Title of the publication. First and corresponding authors were categorized based on author list ordering and correspondence details. Gender was determined by review of the given/first name. For articles that only listed the authors’ first initials, attempts were made to find the full first name through additional publication databases (e.g., PubMed), academic affiliate institutions and internet search engines (e.g., Google). This was manually cross referenced with publications retrieved from research networks in March 2018 and 2020. EndNote™ X8.2 software was used to remove duplicates and update references.

**Table 1.** Search Strategy

|  | *Inclusion Criteria* | *Exclusion Criteria* |
| --- | --- | --- |
| Database | Medline®/PubMed®; Web of Science™ (Thompson Reuters) and Individual Research Networks with a PubMed® unique identifier number | Other databases |
| Publication period | 1994-2019 inclusive | Articles Published in 2020 |
| Research network | RIDEPLA<br>REPEM<br>RISeuP/SPERG<br>PECARN<br>PEM CRC<br>PERC<br>PREDICT<br>PERUKI<br>PERN | Publications without affiliation to PERN |
| Document types | PubMed® identifier number | No PubMed® identifier number |
| Language | All languages | none |

RIDEPLA Red de Investigación y Desarrollo de la Emergencia Pediátrica Latinoamericana; REPEM Research in European Pediatric Emergency Medicine; RISeuP/SPERG Spanish Pediatric Emergency Research Group; PECARN Pediatric Emergency Care Applied Research Network; PEM CRC Pediatric Emergency Medicine Collaborative Research Committee of the American Academy of Pediatrics; PERC Pediatric Emergency Research Canada; PREDICT Paediatric Research in Emergency Departments International Collaborative; PERUKI Paediatric Emergency Research in the United Kingdom and Ireland; PERN Pediatric Emergency Research Network

### Toolkits for Bibliometric analysis

Bibliometric analysis of research network publications includes the year of publication, research areas, types of documents, keywords, language, articles, authors, journals, and countries by PubMed^®^. Citations, citations per year, the number of citations per article and h-index were established in ISI Web of Science. The bibliometric mapping and visualization techniques are based on extensive bibliometric and information visualization literature. ^12^ This was conducted using VOSviewer (Leiden University, Belgium) (http://www.vosviewer.com). ^13^

Terms (Keywords and Authors) are in the 2D map in such a way that the distance between terms gives an indication of their relatedness. The relatedness of terms is determined by counting the number of times terms co-occur. The font size of a term is dependent on the number and strength of co-occurrence with other words in publications. Each of these terms occur a minimum of 5 times. The “overlay visualization” map returns a network that brings incidences of co-authorship or keywords and brings information of chronological order. This visualization is the foundation of the architecture of research that is represented by terms and can be used to assess trends in a research field. ^9^

Publications identified by the search in Table 1 were further analyzed by the Dimensions Artificial Intelligence program (Cambridge, MA, USA) (https://app.dimensions.ai/). ^14^ This links publications and citations with grants, patents, clinical trials, datasets, and policy papers to deliver a more holistic view of the research landscape. Dimensions analysis produces the Altmetric Attention Score, which is a weighted count of the online ‘mentions’ a publication has received. This is a measure of research impact, with immediately available information on the reach and influence of an article, and an ability to track how the attention changes over time. ^15^

### Statistical analysis

Data are presented as median with interquartile range (IQR). Pearson’s correlation coefficient determined associations between continuous variables. Data were analyzed using Microsoft Excel 2016. Statistical significance was set at 2-tailed P <.05.

### Ethical Considerations

This retrospective observational study did not involve human subjects, and ethical approval was not required.

## Results

### Publications

Between 1994 and 2019, a total of 493 publications were published in journals by PEM research networks. The first publication was in 1994, the annual publication count had increased to 97 publications in 2019 (Figure 1), and PERN was established in October 2009 (blue arrow). Since 2004 the number of publications per annum has been steadily increasing. The number of publications per annum has surpassed the linear trendline of annual publications since 2016 (Figure 1). Full free text was available for 250 publications. The gender of the first author was female in 46% (227) of publications. The corresponding author(s) were female in 45% of publications.

**Figure 1.**
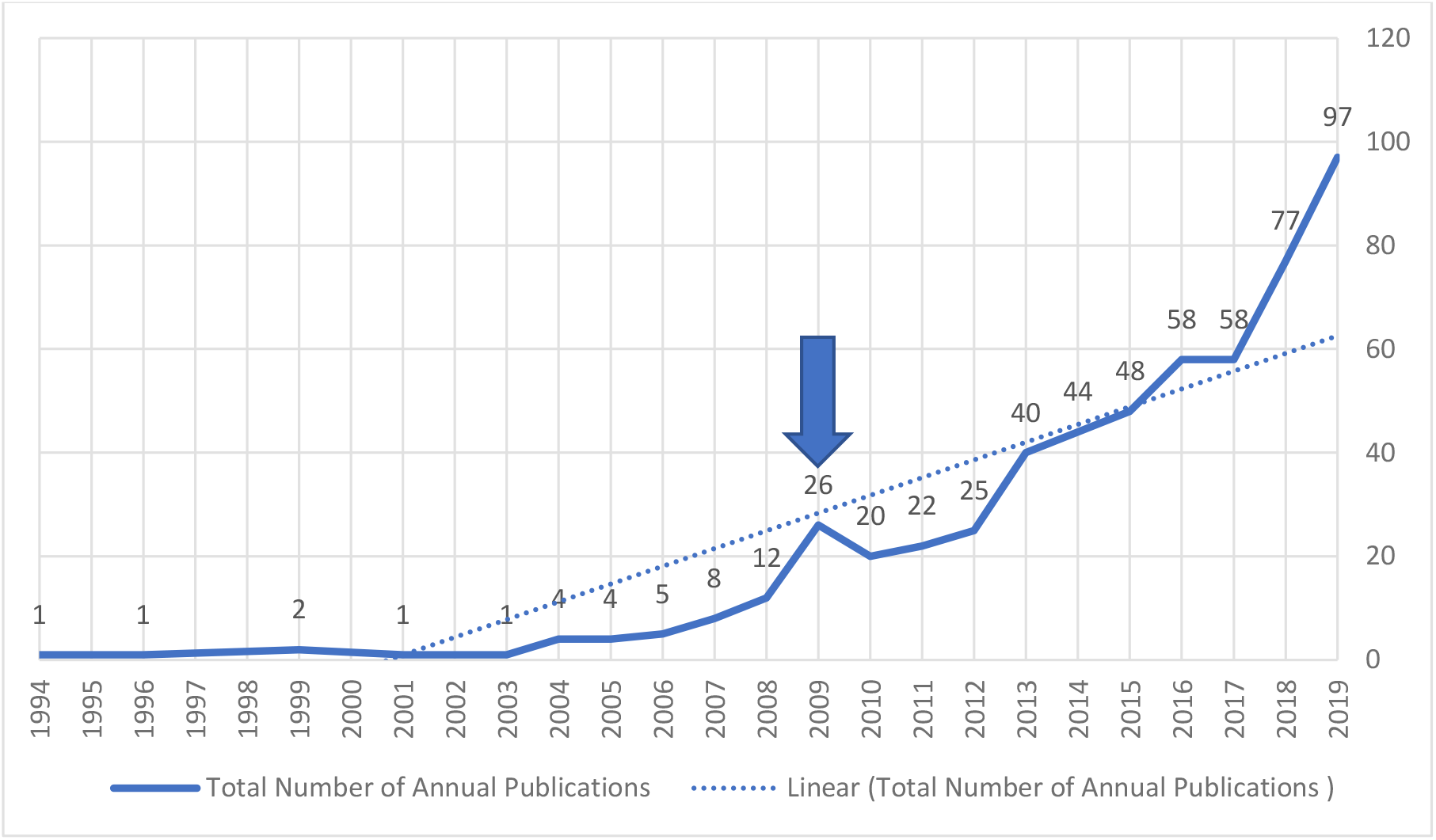
Total annual publications across all networks and number of research networks Blue Arrow: Paediatric Emergency Research Network (PERN) inception in 2009

The range of publication counts by each group and related metrics is demonstrated in Table 2. There was a non-significant moderate positive correlation between the number of years publishing and the number of publications (Pearson’s coefficient R =.58; P=.12). PECARN had the highest number of publications, highest sum of times cited, and h-index, whilst PEM CRC had the highest average citations per publication. PEM CRC is the most mature network and RIDEPLA is the youngest network. A non-significant moderate positive correlation was observed between the number of years publishing and that network’s sum of citations (Pearson’s Correlation Coefficient R =.66; P=.07). There was non-significant moderate negative association between the number of countries in a network and total publications (Pearson’s Correlation Coefficient R = -.53; P=.17).

**Table 2.** Research Network Publication Performance from 1994 to 2019

| Research Network | Number of Countries in Research Network | Number of Years Publishing to 2019 | Publications<sup>a</sup> (average publications per annum of existence) | Sum of Times Cited<sup>o</sup> | Average Citations per item<sup>b</sup> | h-index<sup>b</sup> |
| --- | --- | --- | --- | --- | --- | --- |
| All Networks | 40 | 26 | 493 | 10203 | 20.91 | 48 |
| PECARN | 1 | 15 | 163 (10.8) | 4156 | 26.14 | 33 |
| PEM CRC | 1 | 26 | 47 (1.8) | 1731 | 36.83 | 19 |
| PERC | 1 | 21 | 153 (7.2) | 3225 | 21.22 | 29 |
| PERUKI | 2 | 7 | 27 (3.8) | 158 | 6.08 | 9 |
| PREDICT | 2 | 15 | 108 (7.2) | 1048 | 10.08 | 19 |
| RISeuP.SPERG | 1 | 5 | 13 (2.6) | 39 | 3 | 4 |
| REPEM | 20 | 10 | 11 (2.0) | 106 | 9.64 | 7 |
| RIDEPLA | 14 | 3 | 4 (1.3) | 8 | 2.67 | 2 |
| PERN | 40 | 10 | 12 (1.2) | 82 | 7.45 | 5 |

<sup>a</sup>Duplication exists with Publications shared between Research Networks <sup>b</sup>Derived from ISI Web of Science RIDEPLA Red de Investigacion y Desarrollo de la Emergencia Pediatrica Latinoamericana; REPEM Research in European Pediatric Emergency Medicine; RISeuP/SPERG Spanish Pediatric Emergency Research Group; PECARN Pediatric Emergency Care Applied Research Network; PEM CRC Pediatric Emergency Medicine Collaborative Research Committee of the American Academy of Pediatrics; PERC Pediatric Emergency Research Canada; PREDICT Paediatric Research in Emergency Departments International Collaborative; PERUKI Paediatric Emergency Research in the United Kingdom and Ireland; PERN Pediatric Emergency Research Network

The relatedness of countries and keywords over time reveals initial establishment of the United States (dark green) in the literature with temporal establishment of Canada, Australia, New Zealand (light green) and more recently United Kingdom (and Ireland) and Spain (yellow) (Figure 2a). A similar temporal progression of publication keywords is noted with croup (dark green) progressing sequentially to bronchiolitis/asthma, meningitis, fever, sepsis/resuscitation, pain management, wounds and injuries, athletic injuries and more recently onto traumatic brain injuries (yellow) (Figure 2a).

**Figure 2a.**
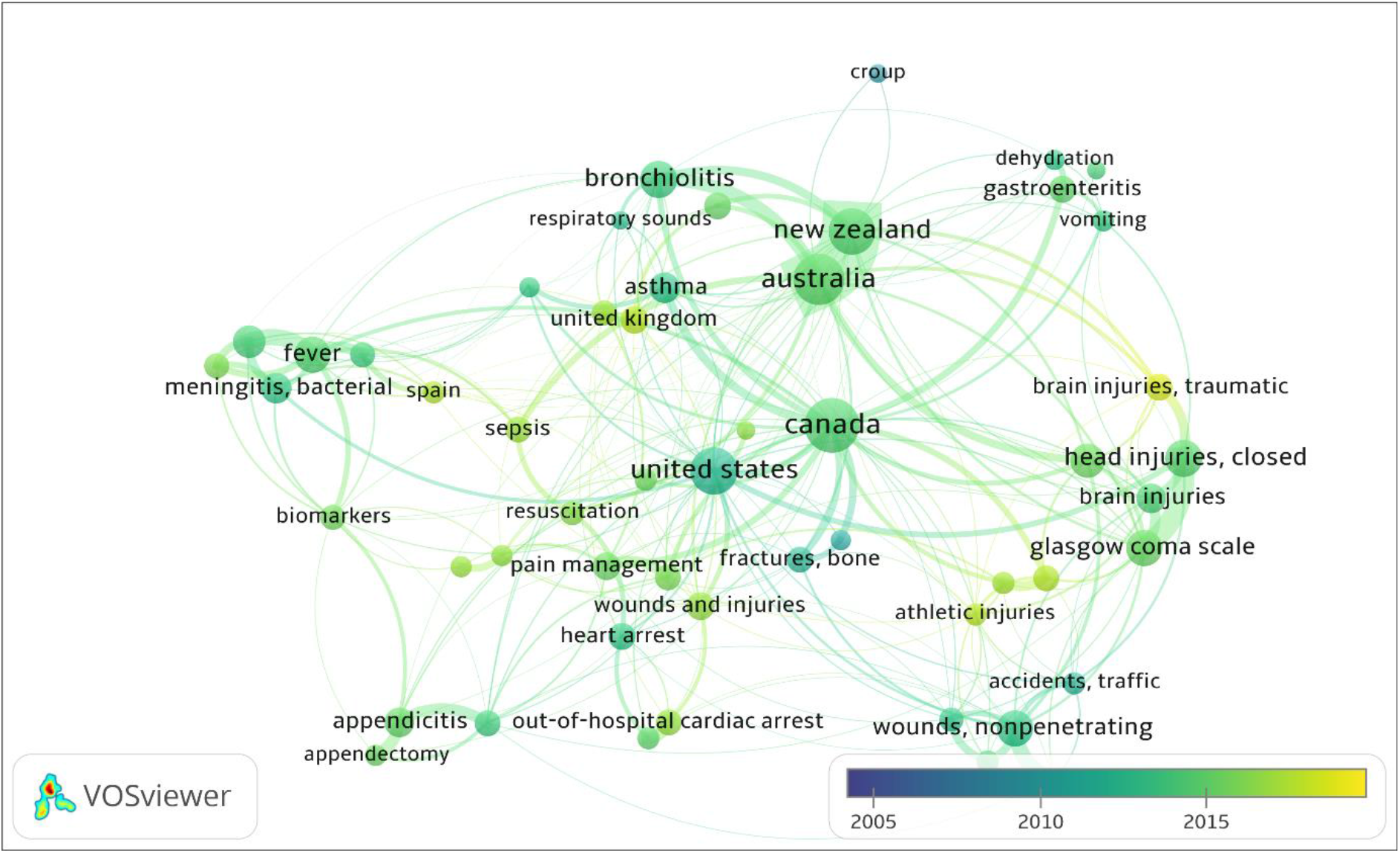
Temporal trends of the regional network output and progression of topics denoted by keywords from 2005-2019

The most productive authors are clearly identified by the size of the weighted circles (Figure 2b) and appear to be centered within distinctive groups of international authors (e.g., Kuppermann/Dayan, Babl/Lyttle/Borland/Bressan/Neutz and Freedman/Plint/Boutis). The multiple and extensive collaborations between authors and groups is clearly demonstrated by lines travelling between authors within and across separate groupings (Figure 1b). The temporal development of authors and their collaborations are clearly demonstrated by the “overlay visualization” in Figure 2b.

**Figure 2b:**
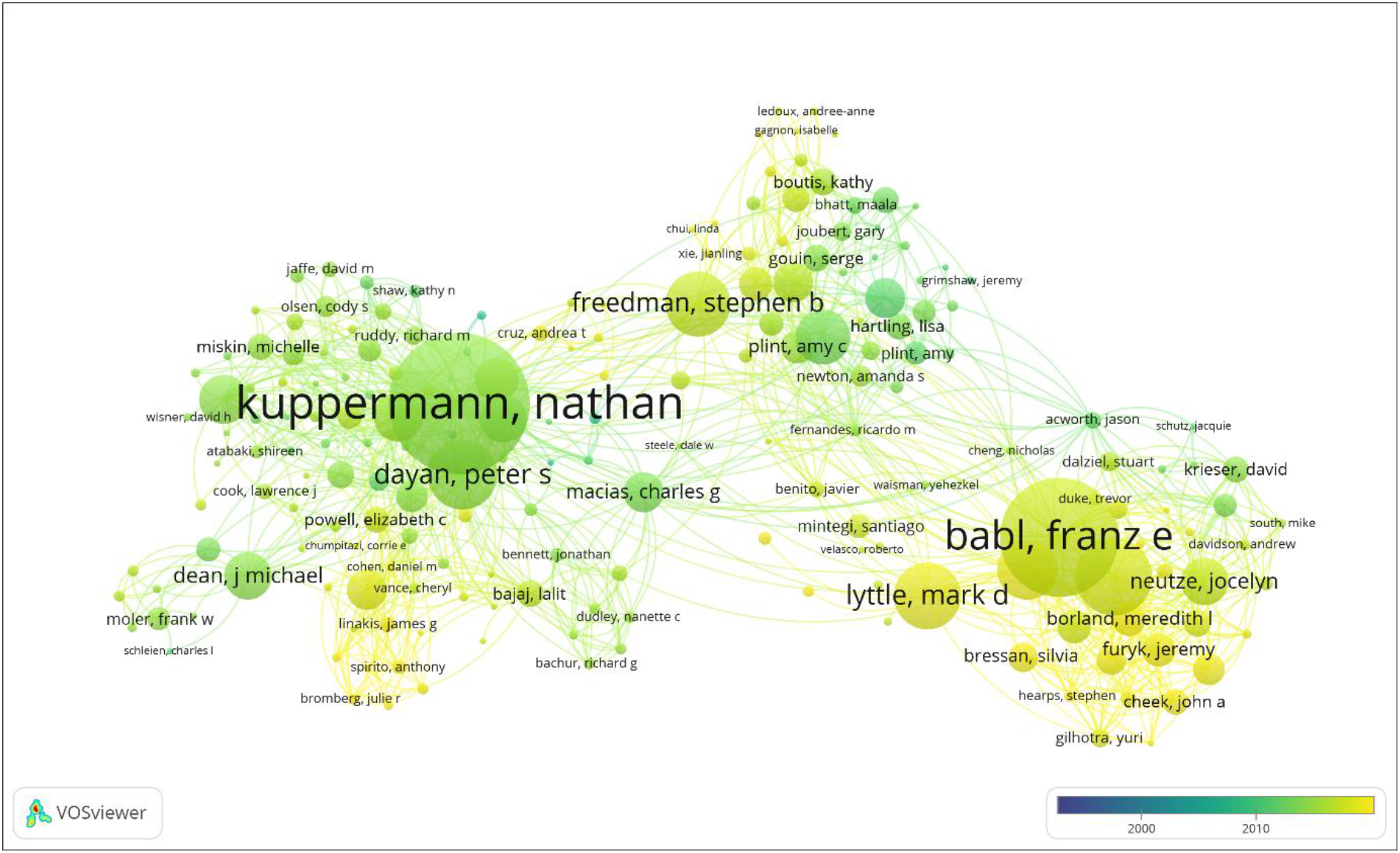
Map of Authors and collaborations in Publications from 1994-2019 Legend: More productive authors are identified by the larger size of the weighted circles and linking lines reveal collaborations

The most impactful research publications have been produced by PECARN, PERC and PEMCRC (Table 3) and these are the 3 longest formed research networks (Table 2). The most impactful publications have all been issued in the last three years. The median (IQR) year of most impactful publications was 2018 (2016-2019). The topics of these articles were concussion, status epilepticus, gastroenteritis, asthma, bronchiolitis, diabetic ketoacidosis, serious bacterial infection, and procedural sedation. Five of the top 10 impactful publications have been published in *JAMA* publications, four in the *New England Journal of Medicine* and one in *Pediatrics*.

**Table 3.** Top 10 Cited and Impactful Publications from Research Networks

| Top 10 Impactful Publications<sup>a</sup> |  | Altmetric Attention Score |  |
| --- | --- | --- | --- |
| 1. | Kapur, Jaideep, et al. "Randomized trial of three anticonvulsant medications for status epilepticus." *New England Journal of Medicine* 381.22 (2019): 2103-2113. | 888 |  |
| 2. | Grool, Anne M., et al. "Association between early participation in physical activity following acute concussion and persistent postconcussive symptoms in children and adolescents." *JAMA* 316.23 (2016): 2504-2514. | 851 |  |
| 3. | Freedman, Stephen B., et al. "Multicenter trial of a combination probiotic for children with gastroenteritis." *New England Journal of Medicine* 379.21 (2018): 2015-2026. | 812 |  |
| 4. | Merckx, Joanna, et al. "Respiratory viruses and treatment failure in children with asthma exacerbation." *Pediatrics* 142.1 (2018): e20174105. | 627 |  |
| 5. | Kuppermann, Nathan, et al. "Clinical trial of fluid infusion rates for pediatric diabetic ketoacidosis." *New England Journal of Medicine* 378.24 (2018): 2275-2287. | 540 |  |
| 6. | Franklin, Donna, et al. "A randomized trial of high-flow oxygen therapy in infants with bronchiolitis." *New England Journal of Medicine* 378.12 (2018): 1121-1131. | 449 |  |
| 7. | Leddy, John J., et al. "Early subthreshold aerobic exercise for sport-related concussion: a randomized clinical trial." *JAMA Pediatrics* 173.4 (2019): 319-325. | 421 |  |
| 8. | Bhatt, Maala, et al. "Risk factors for adverse events in emergency department procedural sedation for children." *JAMA Pediatrics* 171.10 (2017): 957-964. | 370 |  |
| 9. | Zemek, Roger, et al. "Clinical risk score for persistent postconcussion symptoms among children with acute concussion in the ED." *JAMA* 315.10 (2016): 1014-1025. | 336 |  |
| 10. | Kuppermann, Nathan, et al. "A clinical prediction rule to identify febrile infants 60 days and younger at low risk for serious bacterial infections." *JAMA Pediatrics* 173.4 (2019): 342-351. | 298 |  |
| Top 10 Cited Publications<sup>b</sup> |  | Citations | Average citations per Year |
| 1. | Kuppermann, Nathan, et al. "Identification of children at very low risk of clinically-important brain injuries after head trauma: a prospective cohort study." *The Lancet* 374.9696 (2009): 1160-1170. | 739 | 61.58 |
| 2. | Glaser, Nicole, et al. "Risk factors for cerebral edema in children with diabetic ketoacidosis." *New England Journal of Medicine* 344.4 (2001): 264-269. | 398 | 19.9 |
| 3. | Osmond, Martin H., et al. "CATCH: a clinical decision rule for the use of computed tomography in children with minor head injury." *CMAJ* 182.4 (2010): 341-348. | 254 | 23.09 |
| 4. | Moler, Frank W., et al. "Therapeutic hypothermia after out-of-hospital cardiac arrest in children." *New England Journal of Medicine* 372.20 (2015): 1898-1908. | 205 | 34.17 |
| 5. | Zemek, Roger, et al. "Clinical risk score for persistent postconcussion symptoms among children with acute concussion in the ED." *JAMA* 315.10 (2016): 1014-1025. | 184 | 36.8 |
| 6. | Nigrovic, Lise E., et al. "Clinical prediction rule for identifying children with cerebrospinal fluid pleocytosis at very low risk of bacterial meningitis." *JAMA* 297.1 (2007): 52-60. | 184 | 13.14 |
| 7. | Corneli, Howard M., et al. "A multicenter, randomized, controlled trial of dexamethasone for bronchiolitis." *New England Journal of Medicine* 357.4 (2007): 331-339. | 170 | 12.14 |
| 8. | Levine, Deborah A., et al. "Risk of serious bacterial infection in young febrile infants with respiratory syncytial virus infections." *Pediatrics* 113.6 (2004): 1728-1734. | 164 | 9.65 |
| 9. | Plint, Amy C., et al. "Epinephrine and dexamethasone in children with bronchiolitis." *New England Journal of Medicine* 360.20 (2009): 2079-2089. | 143 | 11.92 |
| 10. | Meert, Kathleen L., et al. "Multicenter cohort study of in-hospital pediatric cardiac arrest." *Pediatric Critical Care Medicine* 10.5 (2009): 544. | 139 | 11.58 |

<sup>a</sup> Source: Dimensions https://app.dimensions.ai/ <sup>b</sup> Source: Web of Science

The top cited publications have been produced by PECARN, PERC and PEMCRC (Table 3). These publications have been issued over the past 19 years. The median (IQR) year of top cited publications was 2009 (2006-2011). The topics of these publications were head injury, cardiac arrest, serious bacterial infection, bronchiolitis, and diabetic ketoacidosis. Four were published in the *New England Journal of Medicine*, two in *JAMA*, one each in *The Lancet, Canadian Medical Association Journal, Pediatrics* and *Pediatric Critical Care Medicine*.

## Discussion

This study has examined and compared, through visual and numeric bibliometric analysis, PEM research network publications from 1994 to 2019. The annual product of research networks is increasing, and since the formation of PERN in 2009 the number of global annual publications has increased as has the number of research networks contributing (Figure 1). Five research networks, including PERN, have been established in the last decade, with RIDEPLA the most recently formed in 2016^7^ (Table 2).

Forty six percent of publications had a female first author and 45 % with female corresponding author. This is a much narrower gender gap in first authorship compared to high impact medical journals (37% in 2014) and high impact emergency medicine journals (24-45% in 2018).^16,17^. The most productive networks in terms of number of publications are based in North America, followed by Australia and New Zealand, Europe, and South America. In 2007, Wilson et al reported 58.5% of 14,000 emergency medicine articles were produced from the United States followed by the United Kingdom.^16^ We observed a moderate positive correlation between the number of years publishing and the number of publications. A moderate positive correlation was observed between the number of years publishing and that network’s sum of citations. Interestingly we identified moderate negative association between the number of countries in a network and average publications per annum of existence. Although this relationship is statistically non-significant, this negative correlation may reflect the additional challenges that multinational research networks undertake when conducting studies (e.g., heterogenic capacity to conduct research (i.e., randomized controlled trials), complexity of multi-country governance and research agreements, national legislation, and funding variability). The coalescing of these networks under the banner of PERN has facilitated extensive collaboration between groups of researchers and networks, as is clearly demonstrated in Figure 2b. More mature research networks may potentially provide considerable experiential expertise with respect to the five primary functions of a health research system (1. Governance and management; 2. Financing; 3. Knowledge generation; 4. Utilization and management of knowledge; and 5. Capacity development).^17^ Newer and younger networks potentially stand to gain from this expertise by developing in a shorter duration by learning and collaborating with more established networks.

In bibliometrics current best practice is to use multiple indicators to provide a more robust picture.^11^ We identify the top-cited articles across all networks. All top-cited publications originate out of the 3 networks longest in existence. This may reflect a more mature network’s stage of development with the production of high-quality research. The h-index is a metric to evaluate research productivity and it varies by research field.^18^ In a study of worldwide emergency medicine researchers’ productivity, the h-index had the best coefficient of determination of productivity compared to number of published papers, impact factor or number of citations.^18^ The highest h-index was from the PECARN network closely followed by PERC, PEM CRC and PREDICT (Table 2). The Altmetric Attention Scores can be used to quickly identify which publications have been widely discussed and shared amongst academic and broader audiences.^19^ The top 10 research network publications by Altmetric scores may be one way to discern which articles are deemed important from a dissemination and knowledge translation perspective by assessing non-traditional uptake of the literature. The top scoring articles published help us recognize the quality of the works, discoveries, and trends steering PEM network research.

Our paper has some important limitations. We conducted a visual and numeric analysis of citations and altmetrics from Pediatric Emergency Medicine Research networks affiliated to PERN. To evaluate articles, we had to develop a unique search strategy. We did not provide a dynamic overview of the current state of global PEM research for comparison. However, it is unlikely that this would provide additional material to the global overview that we provide. Publications anecdotally affiliated to a network could not be represented in this study. No one bibliometric measurement is an indicator of the quality of a research group, therefore multiple simple indicators were used to enhance the transparency of the analysis in line with best practice.^11^ We used the Web of Science for our bibliometric analysis. No bibliometric database is superior.^20^ Scopus, Medline, and Google Scholar are other available bibliometric databases. Citation analysis is based on the absolute number of citations that an article receives and many factors that affect citation rates such as older articles. We did not perform an analysis to correlate study funding or study design to research networks.

## Conclusions

The formation of research networks has led to increased volume, as well as increased excellence determined by h-index and citations in PEM research. This study is the first bibliometric analysis of publications from PEM research networks that unite globally and provides insights into the performance of the literature produced. We have applied large-scale data analysis to PEM research network publications using numerical bibliometric indicators and visualizations of productivity. We have identified the gender gap in first authorship compared to high impact medical journals and high impact emergency medicine journals is narrower. Future research to explore these relationships of numerical bibliometric indicators and visualizations of productivity and the temporal trends will benefit the understanding of the generation, reach and dissemination of PEM literature within the global research community.

## Data Availability

All data is available from the corresponding author upon reasonable request.

